# Virtual myocardial blood flow and flow reserve from static PET imaging using artificial intelligence

**DOI:** 10.64898/2026.02.03.26345376

**Authors:** Meghana Urs, Jacek Kwieciński, Mark Lemley, Panithaya Chareonthaitawee, Giselle Ramirez, Aakash Shanbhag, Aditya Killekar, Robert deKemp, Wanda Acampa, Viet T. Le, Steve Mason, Stacey Knight, Rene R. S. Packard, Mouaz Al-Mallah, Daniel S. Berman, Damini Dey, Robert J. H. Miller, Marcelo Di Carli, Piotr J. Slomka

**Author notes:** Authors contributed equally (Meghana Urs, Jacek Kwieciński). **Address for Correspondence**: Piotr Slomka, PhD, Cedars-Sinai Medical Center, 6500 Wilshire Blvd, Los Angeles, California 90048.

## Abstract

**Background:** Quantitative myocardial blood flow (MBF) and myocardial flow reserve (MFR) provide incremental diagnostic and prognostic value in cardiac PET, but their widespread use is limited by the technical demands of dynamic imaging protocols. We evaluated the feasibility of using artificial intelligence (AI) to predict MBF and MFR from static and gated PET images, without the need for dynamic acquisition.

**Methods:** A machine learning (XGBoost) model was trained on 82Rb PET multi-center dataset using static perfusion imaging, injected dose, hemodynamic measures, clinical data and CT-derived features (including body composition) from the hybrid CT attenuation scan. Model performance was evaluated externally in an independent cohort.

**Results:** In total, 10,566 (derivation-cohort) and 7,842 (external-cohort) patients were included in this multi-center study. On the external-cohort, AI approach achieved an Area under the curve (AUC) of 0.92 (0.92–0.93) for abnormal stress MBF and 0.91 (0.90–0.92) for abnormal MFR; Intra-class correlation (ICC) 0.80 (0.78–0.82) and 0.78 (0.76–0.79), respectively. AI MFR closely mirrored the prognostic performance of measured MFR, showing nearly identical Kaplan–Meier risk stratification (both p<0.0001) and maintaining strong, and independently significant associations with all-cause mortality (HR 3.4 [2.8–4.2] vs. 4.6 [3.6–5.8]; both p<0.001), and demonstrated similar added value to perfusion for mortality prediction.

**Conclusion:** AI-predicted virtual stress MBF and MFR assessment using static and gated PET data is feasible and generalizable across cohorts. By removing the dependency on dynamic acquisitions, this approach has the potential to broaden the clinical adoption of flow quantification.

STRUCTURED GRAPHICAL ABSTRACT PET: Positron Emission Tomography, CT: Computed Tomography, MFR: Myocardial Flow Reserve

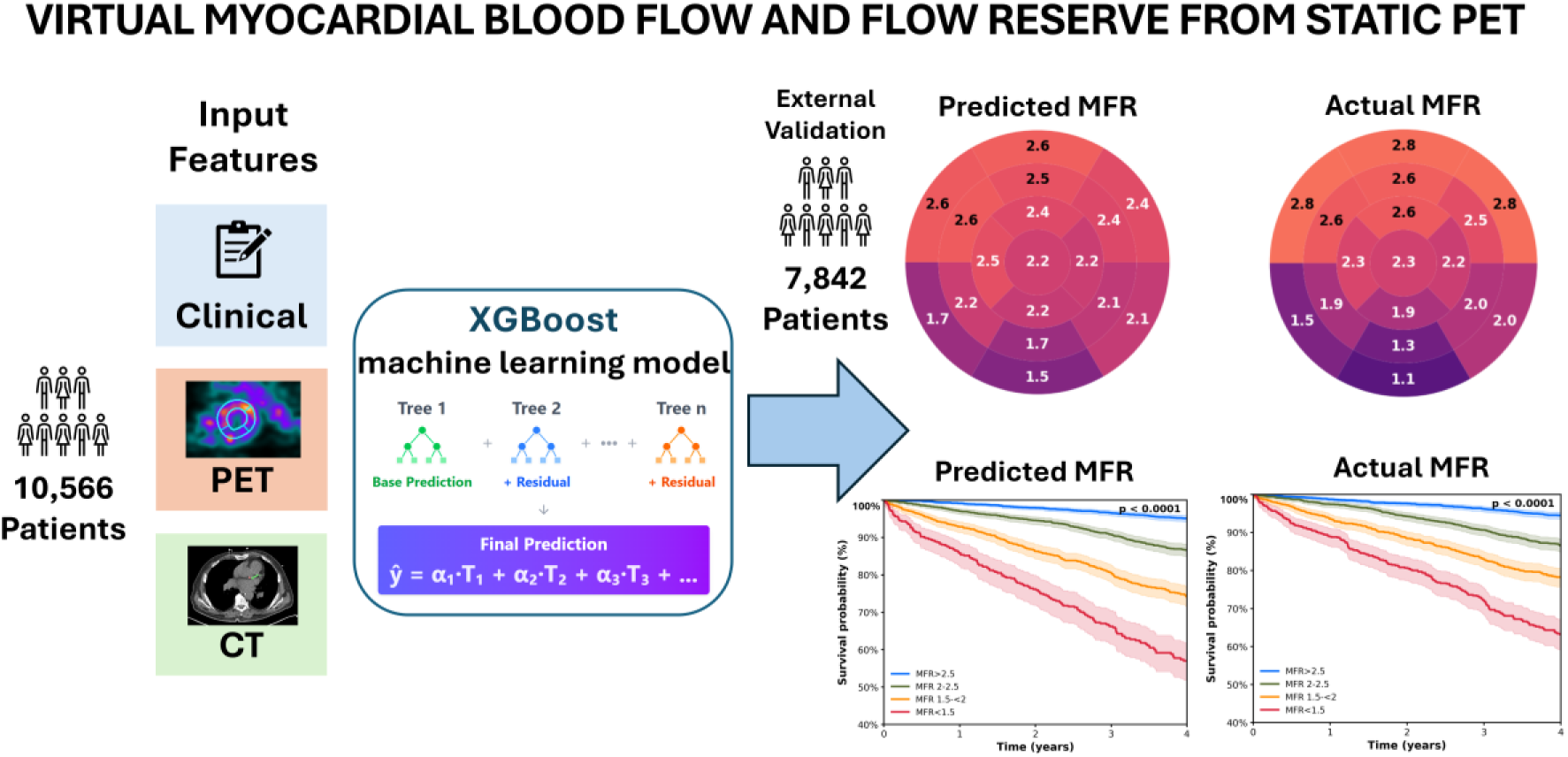

**Key Question:** Can machine learning models trained on dynamic PET datasets accurately predict regional stress myocardial blood flow (MBF) and myocardial flow reserve (MFR) from static image features, physiological parameters, and CT-based anatomical measures?

**Key Finding:** Artificial intelligence can accurately estimate MBF and MFR from non-dynamic PET data, with strong agreement to reference standards.

**Take-home Message:** By eliminating reliance on dynamic PET acquisitions, machine-learning has the potential to broaden clinical adoption of quantitative flow assessment.

## 1. Introduction

Coronary artery disease (CAD) remains the leading cause of mortality and disability worldwide^1^. Myocardial perfusion imaging (MPI) is a cornerstone of CAD evaluation^2^. In the setting of a global epidemic of obesity and diabetes, positron emission tomography (PET), with its ability to quantify myocardial blood flow (MBF) and myocardial flow reserve (MFR), is increasingly employed in the diagnostic work up of CAD^3^ and is recommended by both American and European guidelines for the management of CAD^4–6^.

Despite the proven value of PET derived MBF and MFR as powerful markers of CAD burden and risk stratification, their clinical adoption remains limited. Dynamic PET imaging requires specialized protocols, additional acquisition and reconstruction time, and a high level of technical expertise. As a result, based on Medicare patient data, only a minority of PET centers routinely perform MBF quantification, and data quality may limit reliable implementation^7^.

Emerging artificial intelligence (AI) methods may offer an opportunity to overcome these barriers by predicting virtual MBF and MFR directly from routinely acquired static PET myocardial perfusion studies and auxiliary data. Such an approach could expand the clinical reach of MBF/MFR assessment, particularly in exercise PET protocols enabled by newly approved radiopharmaceuticals (e.g., Flurpiridaz F-18 stress testing), where dynamic acquisitions are logistically impractical^8^, or in cases where the Time activity curves (TACs) are of inadequate quality for conventional MBF measurement.

We hypothesized that machine learning models trained on dynamic PET datasets can accurately predict regional stress MBF and MFR from static image features, physiological parameters, and CT-based anatomical measures – transforming routinely acquired static PET studies into robust quantitative flow assessments without dynamic PET imaging.

## 2. Methods

### 2.1 Patient population

A total of 18,580 patients who underwent clinically indicated dynamic ⁸²Rb PET myocardial perfusion imaging (MPI) were included from six centers^9^—Brigham and Women’s Hospital (BWH), Cedars-Sinai Medical Center (CSMC), University of Naples Federico II, West Los Angeles Veterans Affairs Medical Center (WLAVA), Intermountain Healthcare, and Houston Methodist Academy Institute. Dynamic ⁸²Rb PET served as the reference standard for quantification of stress myocardial blood flow (MBF) and myocardial flow reserve (MFR).

The internal development cohort consisted of patients from CSMC (n = 4,770), BWH (n = 4,187), WLAVA (n = 1,184), and Naples (n = 514). After excluding cases with incomplete data or missing required features, 10,566 of 10,655 patients were used for model training and internal cross-validation. The external validation cohort included independent datasets from two sites: Intermountain Healthcare (n = 6,001) and Houston Methodist (n = 1,924), with 7,842 patients out of 7,925 meeting inclusion criteria for evaluation.

All participating centers obtained institutional review board approval for data use. Oversight of the multicenter study was coordinated through the lead site’s IRB. Each site obtained either written informed consent or a waiver of consent for use of de-identified imaging data, in accordance with local regulatory requirements.

### 2.2 PET Imaging

Rest and stress PET MPI was performed in accordance with clinical guidelines and site-specific protocols. Following acquisition of a low-dose helical CT for attenuation correction, a 6-minute rest list-mode acquisition was initiated immediately before injection of weight-based ^82^Rb (925 to 1850 MBq [25–50 mCi]) for both rest and stress studies. Following pharmacological stress, a 6-minute stress list-mode acquisition was performed.

Semiquantitative expert interpretation was performed during clinical reporting, in accordance with societal guidelines, by experienced board-certified nuclear cardiologists with access to all available data, including stress and rest perfusion, gated functional information, and relevant clinical data. Regional myocardial perfusion was assessed using a 17-segment model and a 5-point scoring system (0=normal to 4=absence of detectable tracer uptake). Summed stress, rest, and difference scores (SSS, SRS, and SDS) were calculated.

Rest and stress MBF and MFR were quantified using a 1-tissue compartment kinetic model (QPET, Cedars-Sinai, Los Angeles, CA)^10^. MBF and from blood-to-myocardium spillover fractions were estimated by numeric optimization. Stress and rest flow values (mL/g/min) were computed on a per-segment basis using polar maps, and MFR was calculated as the ratio of stress over rest MBF. Throughout this manuscript, dynamically derived MBF and MFR are referred to as dynamic MBF and MFR.

Coronary artery calcium (CAC) segmentation and scoring were performed using a previously validated deep learning model^11–13^.

### 2.3 Input features

Static myocardial perfusion data were derived from routine rest–stress PET reconstructions. To augment the predictive modeling, a set of auxiliary features were incorporated. These included procedural variables such as injected tracer dose and patient-specific hemodynamic parameters (heart rate and blood pressure at the time of imaging). Structural information was provided by AI-driven, computed tomography attenuation correction (CTAC) derived anatomical measures^14,15^, including volume, mean and median density (HU) for cardiac chambers, aorta, and pulmonary artery. Physiologic context was captured using body composition metrics^16^ such as muscle volume and attenuation (HU).

Together, these non-dynamic features were integrated with perfusion-derived data, including perfusion defect severity and stress-rest change (%)^17^, to support model training and improve the accuracy of MBF and MFR prediction.

### 2.4 Model development

An XGBoost regressor with regularization was developed to predict continuous values of stress MBF and MFR^18^. This gradient-boosting framework was selected for its ability to accommodate heterogeneous feature sets, mitigate overfitting through built-in shrinkage and tree regularization, and model complex nonlinear relationships between inputs and outcomes. Following model training, post-hoc calibration was applied to align predicted distributions with the dynamic PET–derived reference values, thereby improving agreement and reducing systematic bias.

Two complementary models were implemented for myocardial flow prediction: one designed for global MBF/MFR estimation and another focused on regional, segment-level prediction. Both models used log-transformed targets, grid-search tuning, post-hoc isotonic calibration, and target capping (0.3–3.5 with tanh smoothing). Model 1 (Global Model) generated patient-level global estimates and was evaluated with strict 10-fold cross-validation stratified by clinical flow thresholds. Model 2 (Segmental model) extended prediction to the 17-segment American Heart Association (AHA) model using a stacked segment-level dataset incorporating patient features, segment-specific perfusion metrics, and a segment identifier; cross-validation was performed with all segments from each patient retained within the same fold to prevent data leakage.

This dual-model framework enables comprehensive evaluation of both global and regional myocardial flow, facilitating detailed assessment of myocardial perfusion without dynamic imaging. MBF and MFR derived using this framework are referred to in this paper as AI MBF and MFR.

### 2.5 Clinical Outcome

The primary outcome was abnormal stress MBF and MFR measured by standard dynamic PET. Abnormal stress MBF was defined as <1.8 mL/min/g, consistent with established prognostic thresholds, while abnormal MFR was defined as <2.0, reflecting impaired coronary vasodilator capacity^19^.

The diagnostic performance of the minimal segmental predicted MBF and MFR was tested on a subset of patients (n= 740) who underwent invasive coronary angiography within 180 days of PET imaging and no prior CAD, defined as left main stenosis ≥50% or ≥70% in epicardial arteries^20,21^.

In addition to these perfusion-based endpoints, all-cause mortality was evaluated as a key clinical outcome for PET MPI. Mortality was ascertained through the national death index for US-based sites and administrative databases at University of Naples Federico II.

### 2.6 Statistical Analysis

We evaluated the classification performance of the model for detecting clinically abnormal stress MBF and MFR. Model discrimination was quantified using the area under the receiver operating characteristic curve (AUC), along with sensitivity and specificity at the optimal threshold.

Agreement between AI and reference values for both stress MBF and MFR was assessed using the intraclass correlation coefficient (ICC) to quantify reproducibility across continuous measurements. In addition, Bland–Altman (BA) analyses were constructed to check systematic bias and limits of agreement between predicted and reference-standard values.

Segmental analyses were performed using the 17-segment AHA model^22^. Correlation plots were generated to compare predicted and reference values in each myocardial segment, with per-segment Pearson correlations calculated to assess regional reproducibility. Segmental-level BA analyses were additionally performed to characterize bias and variability across myocardial territories.

To evaluate the clinical validity of AI MFR predictions, time-to-event analyses were conducted comparing measured dynamic MFR values with model-predicted AI MFR values in an independent external validation cohort. Patients were stratified into four risk categories based on MFR thresholds: >2.5 (normal), 2.0-2.5 (mildly reduced), 1.5-2.0 (moderately reduced), and <1.5 (severely reduced), consistent with established prognostic risk categories^23^. Kaplan-Meier (KM) survival curves for all-cause mortality were generated for both measured and AI-predicted MFR values, with differences across groups assessed using the log-rank test. Patients-at-risk tables were constructed showing the number of individuals remaining under observation at yearly intervals.

To quantify the association between MFR categories and mortality risk while accounting for potential confounders, Cox proportional hazards regression analyses were performed. Unadjusted and multivariable-adjusted Cox proportional hazards models were fit to estimate hazard ratios (HR) with 95% confidence intervals (CI) for each MFR category relative to the reference group (MFR >2.5). The adjusted model included the following covariates: age, sex, body mass index, hypertension, diabetes mellitus, dyslipidemia, current smoking status, family history of coronary disease, peripheral vascular disease, rate-pressure product, coronary artery calcium score, resting and stress left ventricular ejection fraction, and total perfusion deficit. Statistical significance was defined as p < 0.05. All analyses were performed using Python (version 3.13.5) with the lifelines package (version 0.30.0) for survival analysis.

## 3. Results

### 3.1 Patient population

Patient characteristics are summarized in Table 1. The internal validation cohort comprised 10,655 patients, with a mean age of 68±12 years and 6,511 (61%) were male. After exclusion of cases with incomplete imaging data, 10,566 patients were included for model training. Of these, 4,042 (38%) had reduced MFR, and 2,721 (26%) had abnormal stress MBF.

**TABLE 1.**
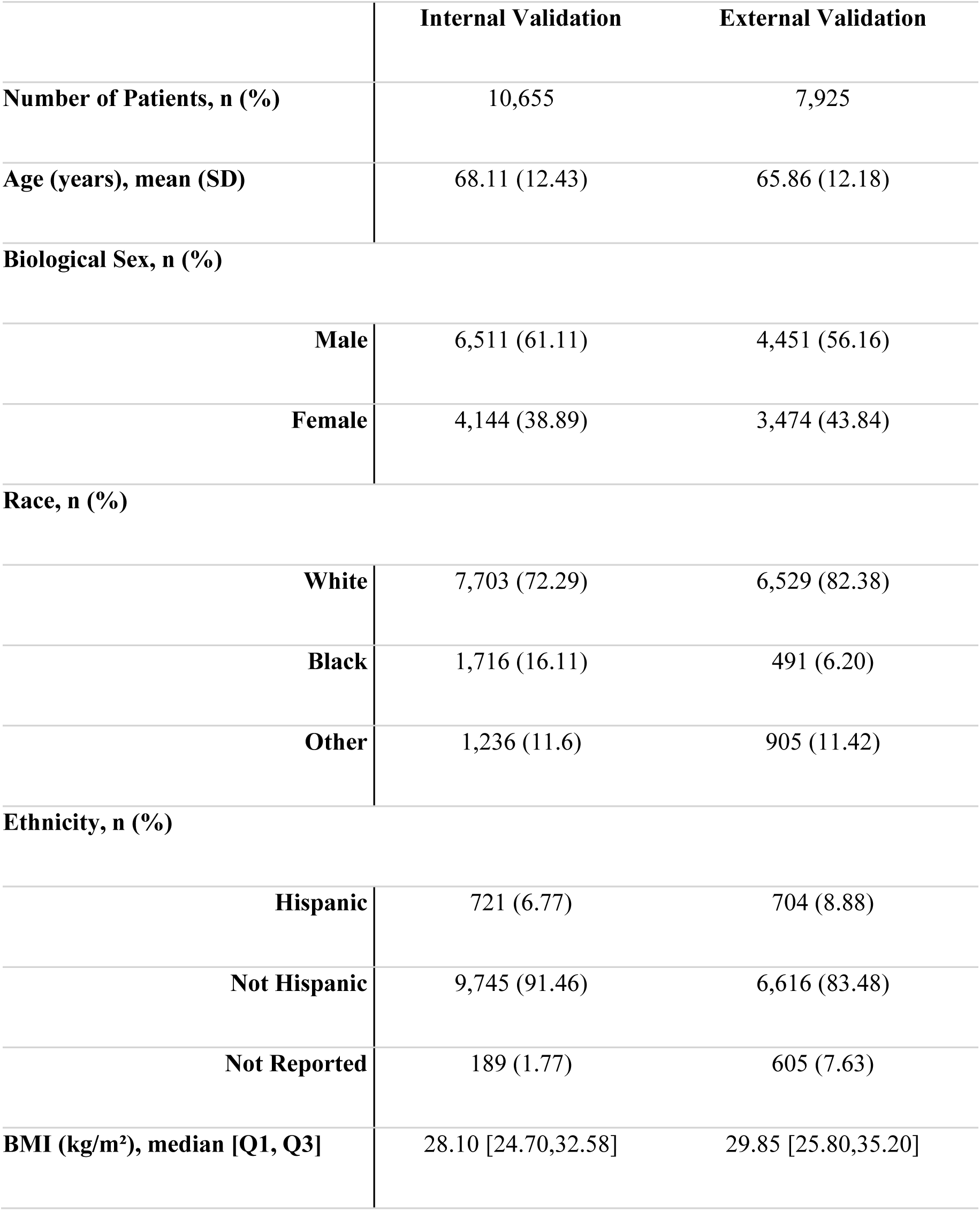
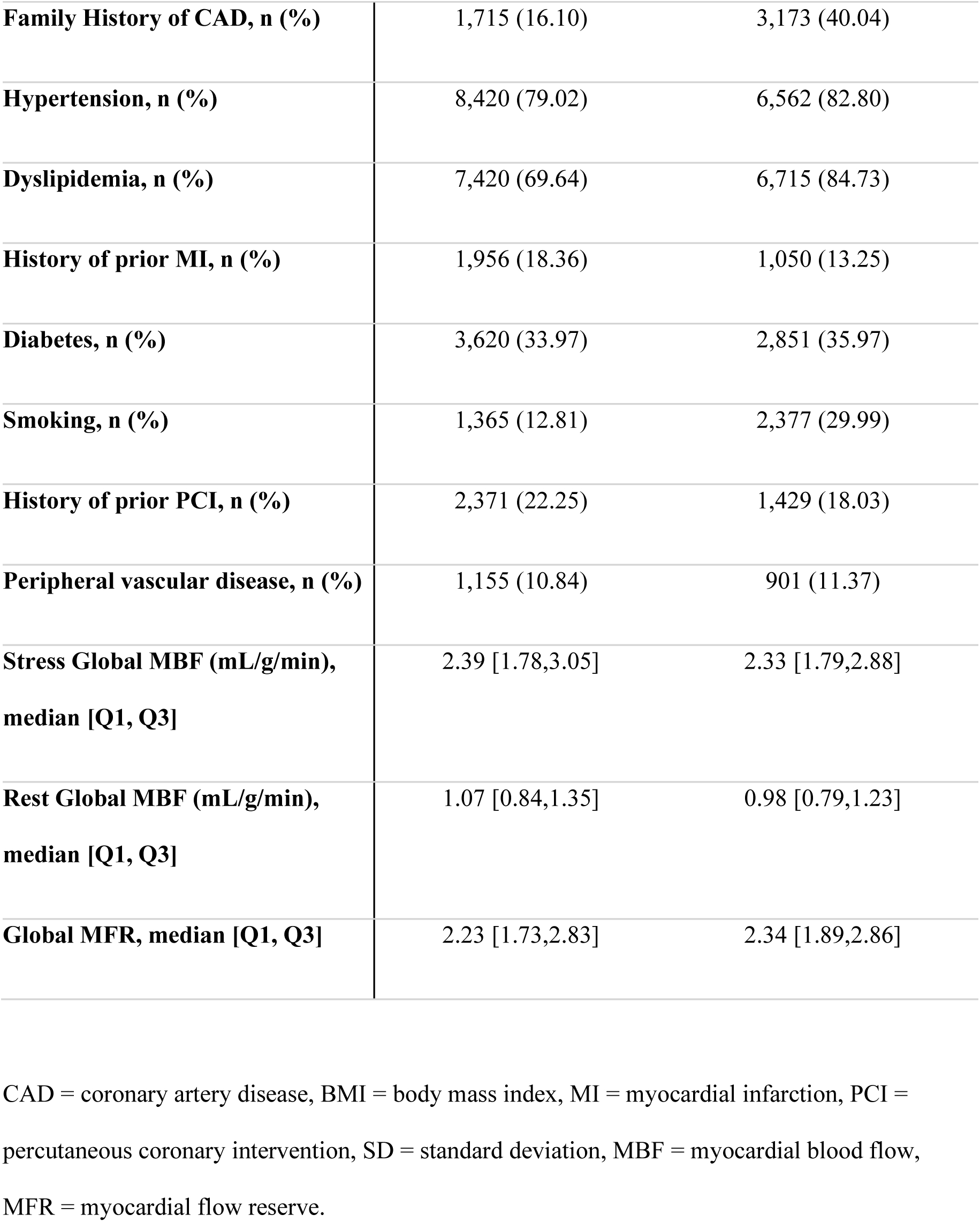
Patient Characteristics.

|  | Internal Validation | External Validation |
| --- | --- | --- |
| **Number of Patients, n (%)** | 10,655 | 7,925 |
| **Age (years), mean (SD)** | 68.11 (12.43) | 65.86 (12.18) |
| **Biological Sex, n (%)** |  |  |
| **Male** | 6,511 (61.11) | 4,451 (56.16) |
| **Female** | 4,144 (38.89) | 3,474 (43.84) |
| **Race, n (%)** |  |  |
| **White** | 7,703 (72.29) | 6,529 (82.38) |
| **Black** | 1,716 (16.11) | 491 (6.20) |
| **Other** | 1,236 (11.6) | 905 (11.42) |
| **Ethnicity, n (%)** |  |  |
| **Hispanic** | 721 (6.77) | 704 (8.88) |
| **Not Hispanic** | 9,745 (91.46) | 6,616 (83.48) |
| **Not Reported** | 189 (1.77) | 605 (7.63) |
| **BMI (kg/m<sup>2</sup>), median [Q1, Q3]** | 28.10 [24.70,32.58] | 29.85 [25.80,35.20] |
| **Family History of CAD, n (%)** | 1,715 (16.10) | 3,173 (40.04) |
| **Hypertension, n (%)** | 8,420 (79.02) | 6,562 (82.80) |
| **Dyslipidemia, n (%)** | 7,420 (69.64) | 6,715 (84.73) |
| **History of prior MI, n (%)** | 1,956 (18.36) | 1,050 (13.25) |
| **Diabetes, n (%)** | 3,620 (33.97) | 2,851 (35.97) |
| **Smoking, n (%)** | 1,365 (12.81) | 2,377 (29.99) |
| **History of prior PCI, n (%)** | 2,371 (22.25) | 1,429 (18.03) |
| **Peripheral vascular disease, n (%)** | 1,155 (10.84) | 901 (11.37) |
| **Stress Global MBF (mL/g/min),**<br>**median [Q1, Q3]** | 2.39 [1.78,3.05] | 2.33 [1.79,2.88] |
| **Rest Global MBF (mL/g/min),**<br>**median [Q1, Q3]** | 1.07 [0.84,1.35] | 0.98 [0.79,1.23] |
| **Global MFR, median [Q1, Q3]** | 2.23 [1.73,2.83] | 2.34 [1.89,2.86] |

CAD = coronary artery disease, BMI = body mass index, MI = myocardial infarction, PCI = percutaneous coronary intervention, SD = standard deviation, MBF = myocardial blood flow, MFR = myocardial flow reserve.

The external validation cohort comprised 7,925 patients, with a mean age of 66±12; 4,451 (56%) were male. Following exclusion of cases with incomplete data, 7,842 patients were included for external validation. Among these, 2,397 (31%) had reduced MFR, and 1,997 (26%) had abnormal stress MBF.

### 3.2 Agreement Analysis

#### Internal Validation

Within the multicenter development cohort, the AI-based approach demonstrated excellent diagnostic performance. For abnormal stress MBF (<1.8 mL/min/g), the model achieved an AUC of 0.91 (95% CI: 0.91–0.92). Similarly, for abnormal MFR (<2.0), the AUC was 0.88 (95% CI: 0.87–0.89). Agreement between predicted and reference continuous values was also very strong, with an ICC of 0.79 (95% CI 0.78-0.8) for stress MBF and 0.73 (95% CI 0.72-0.74) for MFR. Bland–Altman (BA) analysis demonstrated excellent reproducibility, with 95% limits of agreement of ±0.85 for MBF and ±0.90 for MFR, consistent with expected physiologic variability in multicenter ⁸²Rb PET datasets.

#### External Validation

Performance remained consistent when evaluated in two fully independent external cohorts. The model achieved an AUC of 0.92 (95% CI: 0.92–0.93) for detecting abnormal stress MBF and 0.91 (95% CI: 0.90–0.92) for abnormal MFR (Figure 1). Agreement with PET-derived reference values was strong, with ICCs of 0.80 (95% CI: 0.78–0.82) for stress MBF and 0.78 (95% CI: 0.76–0.79) for MFR. BA analysis showed minimal systemic bias (mean bias: –0.09 for MBF; +0.08 for MFR), with 95% limits of agreement of ±0.80 for stress MBF and ±0.78 for MFR, consistent with expected physiologic variability in multicenter ⁸²Rb PET datasets.

**Figure 1.**
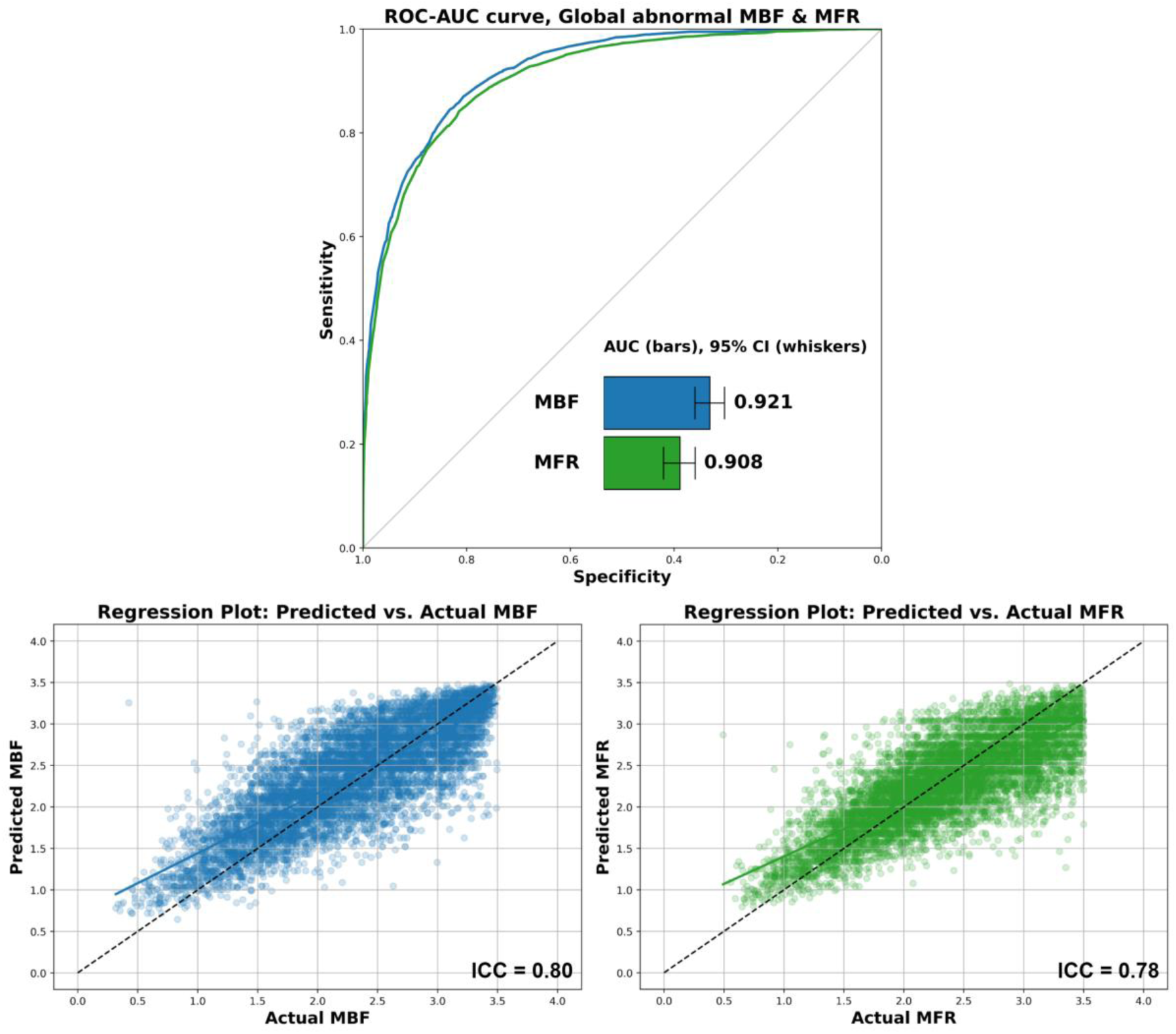
ROC curve for AI detection of abnormal global stress myocardial blood flow (MBF < 1.8) and abnormal myocardial flow reserve (MFR < 2.0) in external dataset (top). Correlation between the actual and virtual MBF and MFR in external dataset (bottom). ROC: Receiver Operating Characteristic, AUC: Area Under the Curve, CI: Confidence Interval, ICC: Intra Class Correlation

### 3.2 Segmental Analysis

The segmental model provided accurate 17-segment MBF and MFR estimates. We observed consistent correlations between actual and AI-predicted MBF values across both internal testing 0.74–0.81 and external validation 0.72-0.82, corresponding to high ICC 0.80 and 0.80 respectively. Concordant findings were found for MFR, with correlations of 0.67-0.74 internally and 0.71-0.77 externally, and ICC 0.69 and 0.73 respectively (Figure 2).

**Figure 2.**
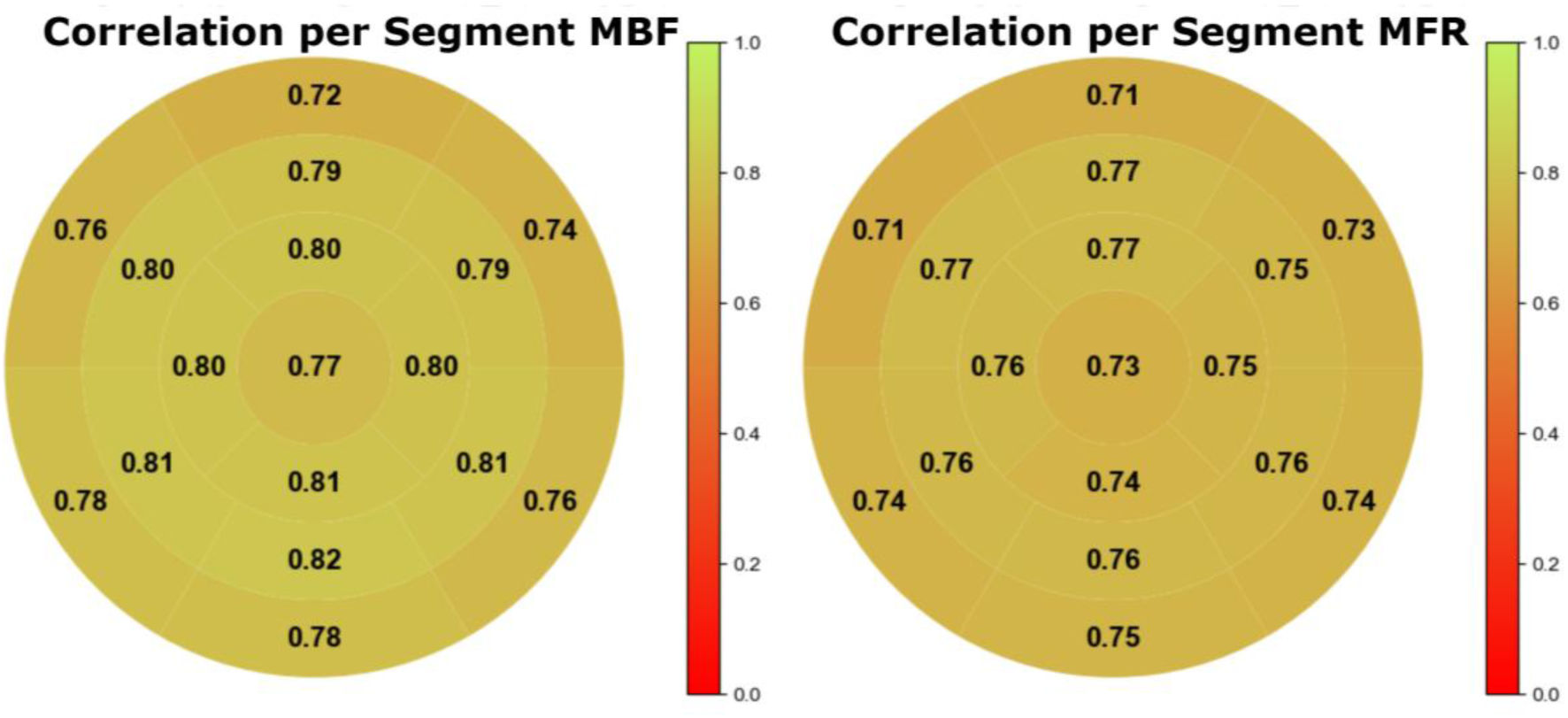
Per-segment Concordance Correlation Coefficients of true and predicted myocardial blood flow (MBF) and myocardial flow reserve (MFR) values for external dataset.

### 3.3 Diagnostic Performance and Clinical outcomes

The accuracy and clinical value of the AI-predicted MBF/MFR was further demonstrated by diagnostic performance and long-term outcome analyses. Among 740 patients in the diagnostic cohort, 345 (46.6%) had CAD on invasive angiography. AI MBF and MFR had an AUC of 0.75 (95% CI: 0.71–0.79) and 0.80 (95% CI: 0.77–0.83), respectively, for the diagnosis of significant CAD, which was numerically similar to those obtained using measured MBF and MFR (AUCs of 0.74 [95% CI: 0.71–0.78] and 0.77 [95% CI: 0.74–0.80]). The difference was not statistically significant for MBF (p = 0.52) but was significant for MFR (p = 0.01), indicating superior diagnostic performance of predicted MFR.

During a median follow-up of 4.3 (IQR 2.8 – 4.9) years, 1,188 (15.1%) patients died. Similar to actual MFR, AI MFR was a strong independent predictor of all-cause mortality with a stepwise increase in mortality across subpopulations of patients stratified according to MFR (Figure 3). Compared to patients with MFR>2.5 those with reduced predicted MFR<1.5 experienced a more than 5-fold increase in mortality risk, comparable to that observed for actual MFR<1.5 (HR 10.3 (95% CI: 8.5-12.4, and HR 7.0 95% CI: 5.9-8.4, respectively; both p < 0.001).

**Figure 3.**
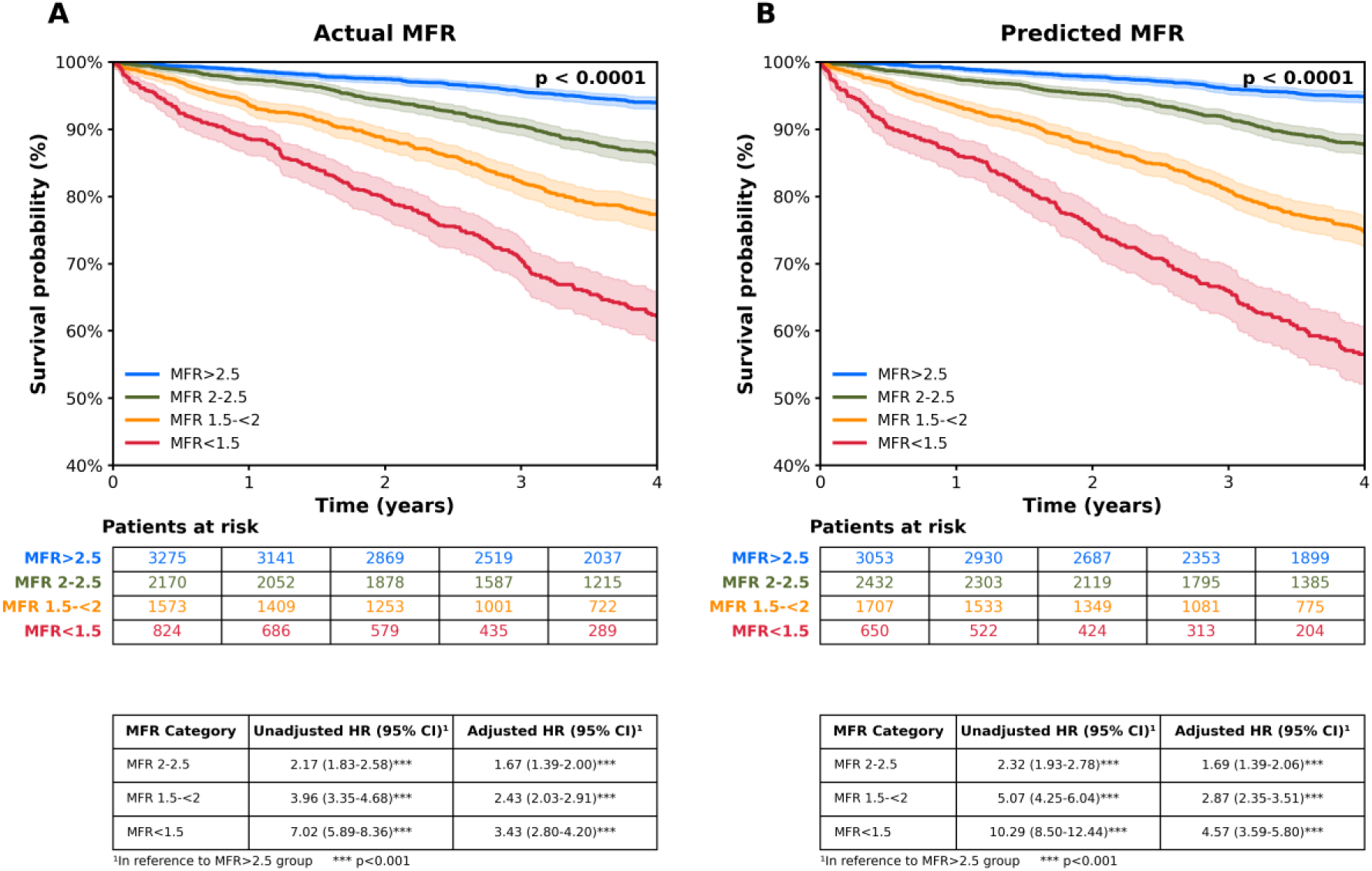
Cumulative incidence of all-cause mortality across actual and predicted myocardial flow reserve (MFR) **in external cohort**. Kaplan-Meier curves for unadjusted survival estimates as a function of true and predicted global MFR for external validation set. Cox proportional hazards regression analyses for actual and predicted MFR, unadjusted and multivariable-adjusted on following co-variates: age, sex, body mass index, hypertension, diabetes mellitus, dyslipidemia, current smoking status, family history of coronary disease, peripheral vascular disease, rate-pressure product, coronary artery calcium score, resting and stress left ventricular ejection fraction. HR: Hazard Ratio, CI: Confidence Interval

In addition, similar to conventional MFR, AI-predicted MFR further risk stratified patients already classified by the presence of perfusion deficits (Figure 4). After adjustments for age, sex, comorbidities, and coronary calcium score, both measured and AI MFR remained strong and independent predictors of all-cause mortality (HR: 3.4 95% CI: 2.8-4.2, vs HR: 4.6 95% CI: 3.6-5.8; both p<0.001).

**Figure 4.**
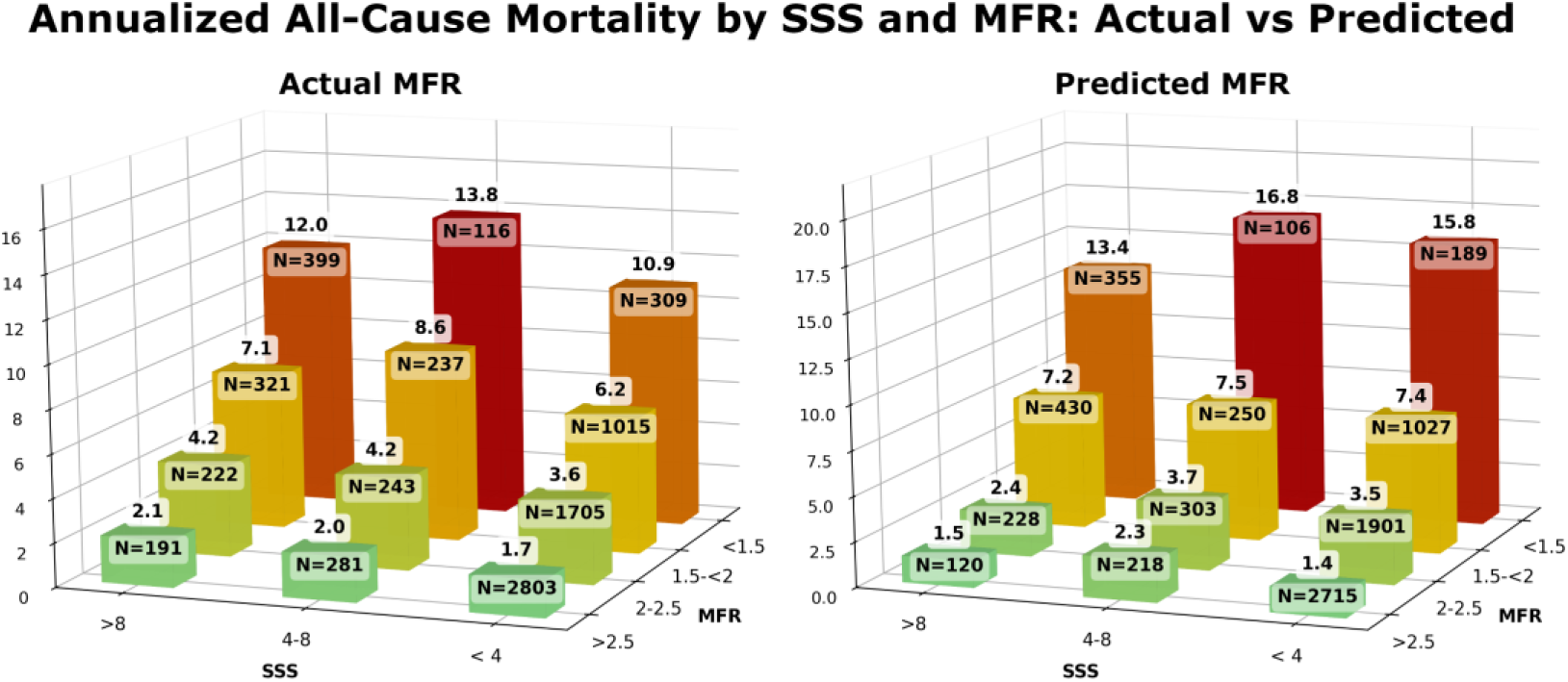
Annualized all-cause mortality according to perfusion and flow reserve for actual versus predicted myocardial flow reserve (MFR) **in external cohort**. The annual rate of cardiac death increased with increasing summed stress score and decreasing MFR. Importantly, lower MFR consistently identified higher-risk patients at every level of myocardial ischemia including among those with visually normal positron emission tomography scans.

### 3.4 Feature importance

Perfusion-derived stress metrics were the dominant contributors in both models (Figure 5), with MFR prediction driven mainly by stress/rest perfusion ratios and MBF prediction influenced by stress ejection fraction and other functional and demographic features.

**Figure 5.**
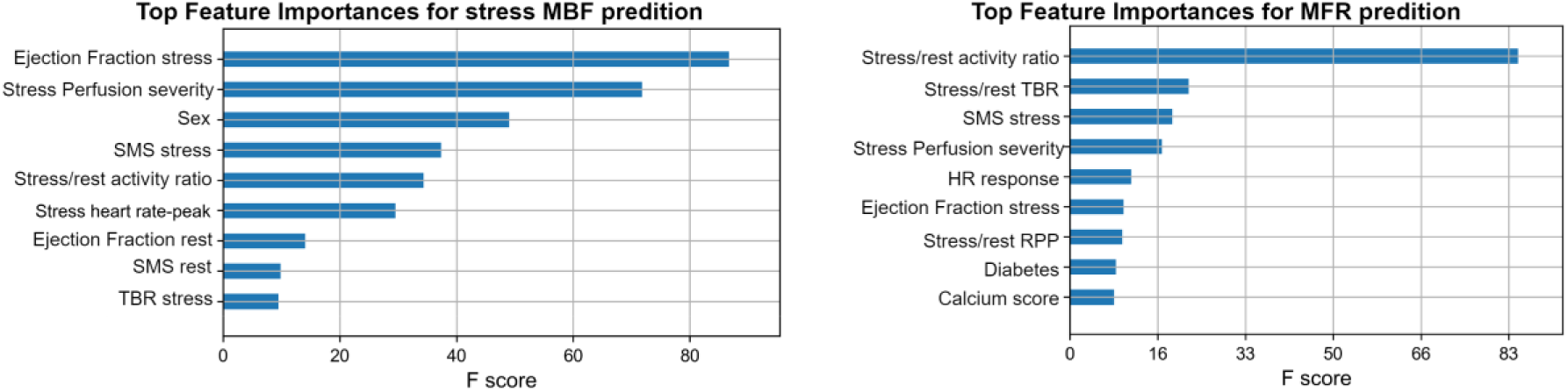
Feature Importance for stress myocardial blood flow (MBF) and Myocardial flow reserve (MFR) prediction. TBR: Target-to-Background Ratio, SMS: Summed Motion Score, HR: Heart Rate, RPP: Rate Pressure Product, BP: Blood Pressure. RCA: Right Coronary Artery

## 4. Discussion

In this study, leveraging state-of-the-art AI, we developed and validated a novel approach for estimating MBF and MFR without the need for dynamic PET acquisitions. This approach represents a fundamental shift in how quantitative PET flow assessment may be performed, enabling robust flow estimation directly from routinely acquired static PET myocardial perfusion studies. Across large, multicenter internal and external cohorts, AI-predicted MBF and MFR demonstrated excellent agreement with dynamic PET reference values, supporting technical validity and scalability of this method.

Collectively, our findings demonstrate that AI-derived flow estimates preserve the essential physiology behavior of PET-based MBF and MFR across a broad range of flow states. The model maintained stability within clinically meaningful flow ranges for diagnosis and risk stratification, while expected variability at higher flow values mirrored known properties of dynamic PET measurements. Importantly, minimal systematic bias and the consistency of performance across independent cohorts support the robustness of this approach and suggest that reliable flow estimation can be effectively recovered using data-driven approaches without direct dynamic acquisition. Together, these observations indicate that the proposed framework captures core flow-related information embedded within routine static PET data, enabling quantitative assessment consistent with established physiologic and clinical paradigms.

Quantitative assessment of MBF and MFR is central to the unique clinical value of PET-MPI^24–28^, and our findings demonstrate that this value can be preserved even in the absence of dynamic acquisition. By enabling accurate estimation of MBF and MFR from non-dynamic PET data, the proposed AI framework retains the key diagnostic strengths of quantitative PET, including reliable identification of hypoperfused myocardium across the full spectrum of disease severity. This capability is particularly relevant for the detection of diffuse or balanced ischemia, where relative perfusion imaging may underestimate disease burden^29^. By extending access to quantitative flow assessment without requiring dynamic imaging protocols, this approach has the potential to broaden the clinical applicability of PET-MPI while maintaining its established physiologic and diagnostic advantages.

Beyond diagnosis, quantitative MFR provides powerful prognostic information, and our results indicate that AI-derived MFR preserves this critical risk stratification capability (Figures 3 and 4). In long-term follow-up, AI-predicted MFR demonstrated a stepwise association with all-cause mortality that closely paralleled measured MFR (Figure 3), including marked risk separation among patients with severely reduced flow. Moreover, consistent with prior seminal observations^23^, AI-derived MFR provided incremental prognostic value beyond relative perfusion abnormalities (Figure 4), reproducing the well-established ability of dynamically derived MFR to refine risk assessment across diverse patient subgroups. These findings reinforce that the clinical utility of MFR lies not only in its absolute value, but in its ability to reclassify risk, an attribute that our AI-based approach appears to retain.

Despite the well-established diagnostic and prognostic value of dynamic PET MBF and MFR replicated across centers and supported by extensive clinical evidence, 2022 Medicare patient data demonstrated that only 39% of sites performing cardiac PET in the USA routinely assessed MBF and MFR^7^. Although multiple factors likely contribute to this underutilization, the technical complexity of acquiring and accurately processing early dynamic data remains a major barrier^30^.

By deriving MBF and MFR from routinely acquired static PET data, the present study directly addresses this limitation and offers a pragmatic pathway to expand access to quantitative flow assessment. This approach has the potential to democratize advanced cardiac PET by enabling flow quantification without the need for protocol modifications, additional acquisition time, or dissemination of specialized technical expertise. Importantly, the level of the agreement between AI-derived and reference dynamic MFR values was numerically comparable to reported scan – rescan reproducibility for ^82^Rb PET (ICCs respectively 0.80 and 0.84) and exceeded the correlation observed between MFR estimates obtained with different PET tracers (^13^N-ammonia versus ^82^Rb PET; R^2^ = 0.633)^31^. Together, these findings suggest that clinically reliable flow information can be recovered from static PET studies with performance approaching the intrinsic reproducibility limits of current PET techniques. By coupling quantitative flow assessment from dynamic acquisition, this framework may fundamentally reshape clinical PET workflows, enabling routine MBF and MFR assessment in settings where dynamic imaging is impractical, such as exercise PET protocols and emerging F-18 tracer-based workflows, thereby expanding the reach of quantitative PET into everyday clinical practice.

The extension of our framework to segmental myocardial analysis demonstrated strong per-segment agreement between AI-derived and dynamic MBF and MFR values, demonstrating the potential of AI-derived flow estimates to inform clinical decision-making analogous to dynamically derived quantitative PET (Figure 2). Physiologic concordance was observed across basal, mid, and apical regions, with the highest agreement observed in mid-ventricular territories. As expected, basal and apical segments exhibited modestly greater variability, consistent with known technical challenges such as motion artifacts, partial volume effects, and thinner myocardial wall geometry. Importantly, no systematic over– or underestimation was observed across segments, indicating unbiased performance despite expected regional variability. The ability to capture segmental flow patterns is particularly relevant for clinical decision-making, where the spatial distribution and heterogeneity of ischemia frequently guide management decisions. This represents a meaningful advance beyond global flow prediction, positioning the framework to support future polar map–based or territory-specific applications. Considering how automatized software is already facilitating image analysis, our AI derived flow values could be easily displayed along standard polar maps supporting physicians at the point of care (Figure 6).

**Figure 6.**
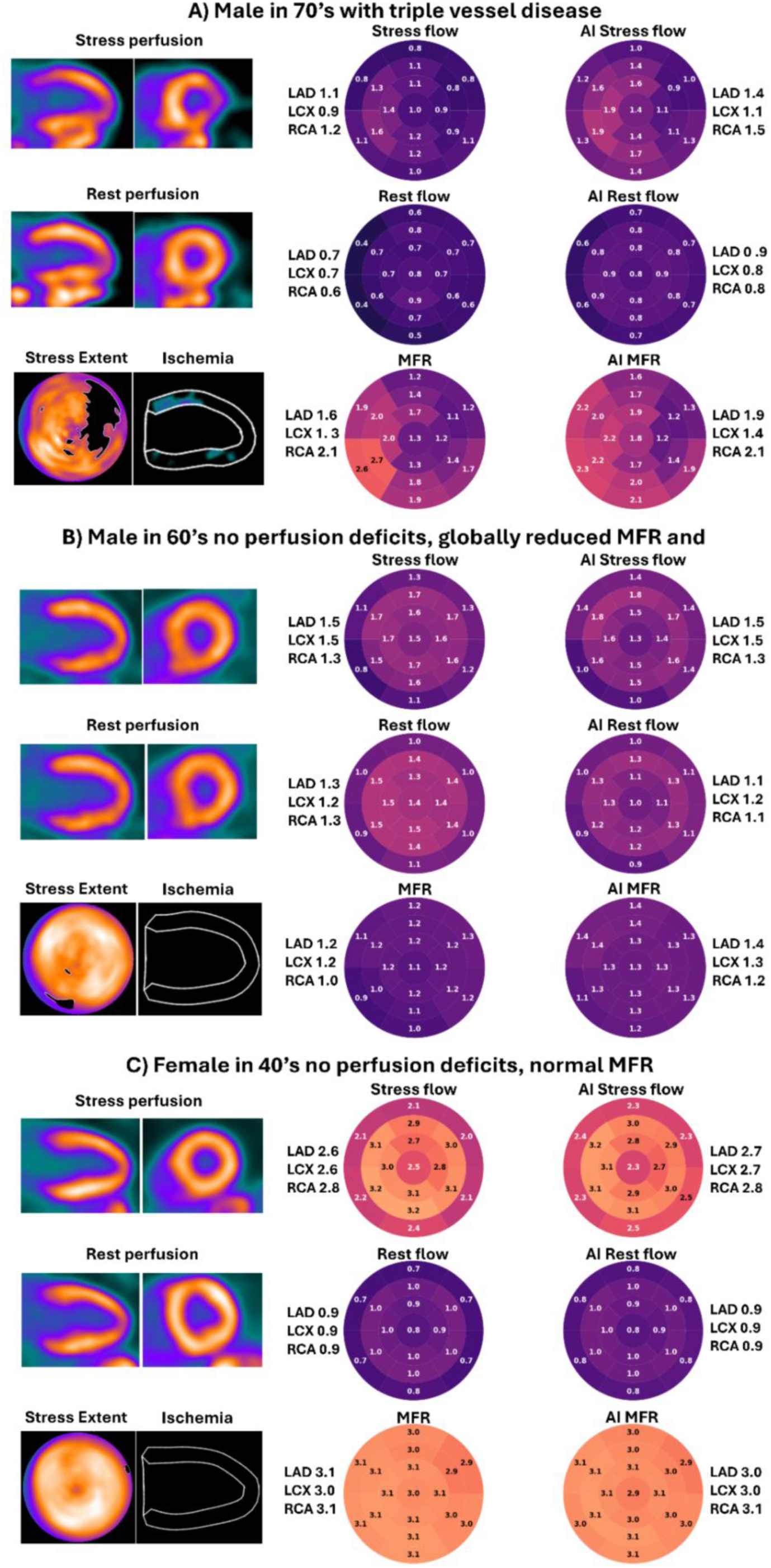
Case examples of actual and virtual MFR. Per-segment dynamic PET–derived reference and AI-predicted virtual stress myocardial blood flow (MBF), rest MBF and myocardial flow reserve (MFR) values. Case A) Male patient in his 70’s with triple vessel disease. Case B) globally reduced MFR in male in his 60’s with no perfusion deficits and death during follow-up. Case C) female in her 40’s, no perfusion deficits and normal MFR.

Examination of feature importance indicated that the models based their predictions on clinically and physiologically meaningful inputs rather than spurious correlations. Stress perfusion-derived features emerged as the dominant contributors, with AI-derived MFR driven primarily by relative stress-to-rest metrics, while AI-MBF incorporated additional influence from functional parameters such as stress ejection fraction and heart rate.

Finally, the goal of this work is not to replace clinician judgment or establish quantitative PET methodologies, but to augment clinical decision-making by expanding access to flow information that is currently underutilized. The proposed AI framework does not operate as a black-box substitute for interpretation; rather, it extracts and synthesizes flow-related information already embedded within routinely acquired PET data and presents it in a familiar, interpretable format. In this context, AI MBF and AI MFR should be viewed as complementary tools that support, rather than supplant, physician expertise – analogous to automated perfusion quantification, gated functional analysis, or coronary calcium scoring that are now integrated into clinical workflows. By reducing technical barriers while preserving physiologic interpretability, this approach aims to enable more consistent and informed use of quantitative PET flow assessment in everyday clinical practice.

### Limitations and Future Directions

This work has several limitations. First, the models were trained and validated using ^82^Rb, and performance with other tracers such as ^13^N-ammonia and ^18^F-flurpiridaz remains to be established. In addition, the present work focuses on prediction of global and segmental MBF and MFR values using tabular data. Future research should aim to extend these methods toward generation of fully image-based flow estimation, which may provide more detailed insights into spatial heterogeneity of perfusion and ischemia. Lastly, this approach could potentially be extended to single photon emission computed tomography and dedicated studies are warranted.

## 5. Conclusion

In summary, this study demonstrates that AI can accurately estimate MBF and MFR from non-dynamic PET data, with strong agreement to reference standards and robust performance across sites. By eliminating reliance on dynamic acquisitions, this approach has the potential to broaden clinical adoption of quantitative flow assessment.

## Data Availability

To the extent allowed by data sharing agreements and IRB protocols, the deidentified data and data analysis code from this manuscript will be shared upon written request.

